# Transient dynamics of SARS-CoV-2 as England exited national lockdown

**DOI:** 10.1101/2020.08.05.20169078

**Authors:** REACT Study Investigators, Steven Riley, Kylie E. C. Ainslie, Oliver Eales, Caroline E. Walters, Haowei Wang, Christina Atchison, Peter J. Diggle, Deborah Ashby, Christl A. Donnelly, Graham Cooke, Wendy Barclay, Helen Ward, Ara Darzi, Paul Elliott

## Abstract

Control of the COVID-19 pandemic requires a detailed understanding of prevalence of SARS-CoV-2 virus in the population. Case-based surveillance is necessarily biased towards symptomatic individuals and sensitive to varying patterns of reporting in space and time. The real-time assessment of community transmission antigen study (REACT-1) is designed to overcome these limitations by obtaining prevalence data based on a nose and throat swab RT-PCR test among a representative community-based sample in England, including asymptomatic individuals. Here, we describe results comparing rounds 1 and 2 carried out during May and mid June / early July 2020 respectively across 315 lower tier local authority areas. In round 1 we found 159 positive samples from 120,620 tested swabs while round 2 there were 123 positive samples from 159,199 tested swabs, indicating a downwards trend in prevalence from 0.13% (95% CI, 0.11%, 0.15%) to 0.077% (0.065%, 0.092%), a halving time of 38 (28, 58) days, and an R of 0.89 (0.86, 0.93). The proportion of swab-positive participants who were asymptomatic at the time of sampling increased from 69% (61%, 76%) in round 1 to 81% (73%, 87%) in round 2. Although health care and care home workers were infected far more frequently than other workers in round 1, the odds were markedly reduced in round 2. Age patterns of infection changed between rounds, with a reduction by a factor of five in prevalence in 18 to 24 year olds. Our data were suggestive of increased risk of infection in Black and Asian (mainly South Asian) ethnicities. Using regional and detailed case location data, we detected increased infection intensity in and near London. Under multiple sensitivity analyses, our results were robust to the possibility of false positives. At the end of the initial lockdown in England, we found continued decline in prevalence and a shift in the pattern of infection by age and occupation. Community-based sampling, including asymptomatic individuals, is necessary to fully understand the nature of ongoing transmission.

## Introduction

SARS-CoV-2 virus has caused high levels of morbidity and mortality globally since emerging in late 2019 (*1*). In response, many populations have greatly reduced levels of social contact either through spontaneous changes in behaviour (*2*) or via government mandated lockdown (*3*). Lockdowns in some countries have been successful in preventing health care systems from becoming overloaded and have brought about reduced rates of virologically confirmed clinical cases (*4*). However this policy has also caused substantial societal and economic harms (*5*) and cannot be maintained indefinitely (*6*).

Full lockdown in the UK was instituted on 23rd March (*7*) after which there was a peak in estimated new infections (*8*) followed by sustained reductions in cases, hospital admissions and deaths (*9*). In order to balance the harms of the virus with those of the lockdown, the UK government began to relax social distancing in England at the end of May. Case incidence continued to fall until 4th July (*10*).

## Methods

To assess the impact of lockdown and its subsequent relaxation -- independent of testing capacity and care-seeking behaviour -- we conducted two rounds of a representative community-based prevalence survey. The survey was designed to provide balanced data for the 315 lower tier local authorities in England. More details on the design and analyses used here are given in (*11*).

Briefly, individuals aged five years and older were randomly selected from the National Health Service (NHS) list of patients registered with a general practitioner and sent an invitation to participate by post. Those who agreed to take part were sent a swab kit and detailed instructions. Parents were asked to conduct the swab for children aged five to twelve years. Personal, demographic and symptom data were obtained via online questionnaire or by telephone. Initially in round 1, wet swabs were sent at ambient temperature to laboratories in the Public Health England (PHE) network (n = 8,595). However, the majority of samples in round 1 (n=112,025) and all samples in round 2 were taken as dry swabs and sent via a temperature controlled cold chain to a commercial laboratory. All swabs were tested for the presence of SARS-CoV-2 virus using RT-PCR. Samples from PHE labs were reported as either positive or negative. All samples from the commercial laboratory for which either the E- or N-gene targets were detected were reported together with gene specific cycle threshold (CT) values.

Cycle threshold values are gene specific and are inversely correlated with the amount of RNA present in a sample. The commercial laboratory reported CT values for all PCRs that detected either the E or N genes, regardless of CT value with samples considered negative when neither E nor N genes were detected. Samples were considered positive when both E and N are detected, regardless of CT values. N was detected in all samples when E was detected. However, in some samples, N was detected but E was not. Based on our earlier work (*11*), samples were called positive with a CT value for N below 37 and called negative with a CT value for N equal to or greater than 37.

We also considered the possible impact of false positive samples on our results driven by our choice of upper CT value for some positive samples. There were differences in the distribution of CT values for the different types of positive results between rounds (Figure S1). Despite a substantial difference in the overall number of samples (Table S1), the percentage of all negatives that had a detected CT value for N gene greater than 37 was the same in both rounds: 0.40% (95% CI, 0.36%, 0.44%) in round 1 and 0.40% (0.37%, 0.43%) in round 2. The percentage of all positives that were positives with CT value for N gene less than 37 was also similar in both rounds: 52% (44%, 60%) in round 1 and 47% (38%, 56%) in round 2. These consistent ratios suggest that the test characteristics were consistent between rounds.

The first round of sample collection was conducted between 1st May and 1st June and obtained 159 positive swabs from 120,610 tested samples (*11*). Here, we present results from the second round of the study (conducted between 19th June and 8th July) and use them with the first round to calculate changes in prevalence, growth rate and reproduction number R. We also analysed key individual characteristics, regional patterns and clustering.

## Results and Discussion

Prevalence of swab-positivity in England fell during the period of our study, from 0.13% (0.11%, 0.15%) in round 1 (*11*) to 0.077% (0.065%, 0.092%) in round 2 (Figure 1A). The decline was not rapid and, based on an exponential decay model, over both rounds combined, corresponded to an average halving time of 38 (28, 58) days, giving a reproduction number R of 0.89 (0.86, 0.93) [assuming an Erlang-distributed serial interval with mean 6 days and shape 2 (*12*)].

**Figure 1.**
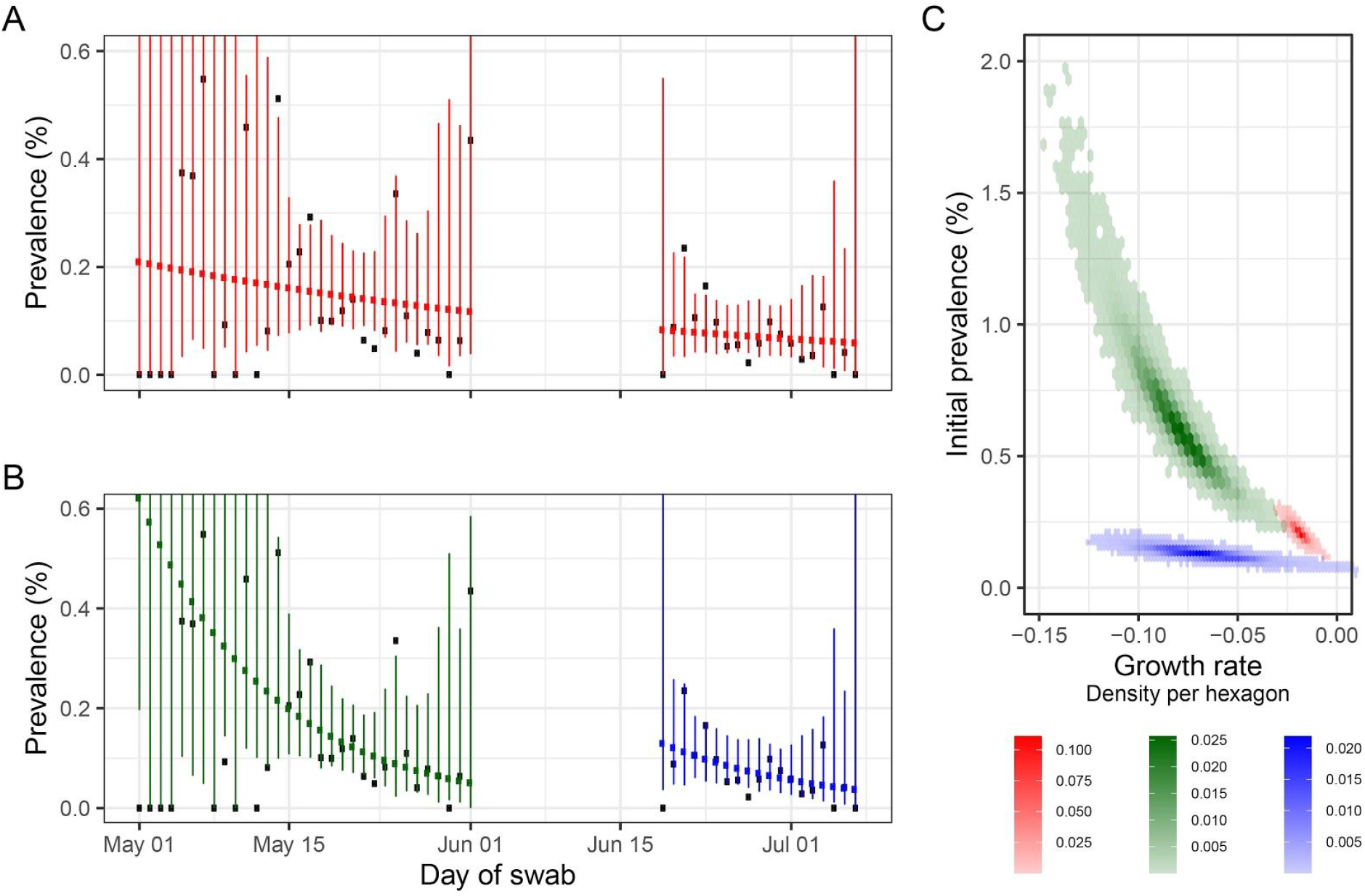
Swab positivity by day with fitted exponential decay models. **A** Observed daily prevalence of swab-positivity for rounds 1 and 2 with an exponential model of decay fit to both rounds as a single dataset. Red shows best fit for constant decay rate (points) with 95% prediction intervals (vertical lines). **B** as A, with models fit separately to rounds 1 and 2. **C** Bivariate posterior density for the three decay models shown in A: red, fit to rounds 1 and 2 (as shown in A); green, fit to only to round 1 (as shown in B, LHS); blue, single round fit to round 2 (as shown in B, RHS).

Although the study was designed to measure a constant decline between sequential rounds (as above). However, we tested the possibility that our data might be better explained by allowing the rate of decline to vary over time: we used a regression framework to fit a smooth term to prevalence and found it was better supported than a constant decline (Figure S2, Table S2). Therefore, we also estimated growth rates and R values for rounds 1 and 2 separately (Figure 1B). Considering each round separately gave a more rapid decline than was estimated using a single growth rate for both rounds For round 1 halving time was 8.6 (6.2, 13.6) days and R was 0.57 (0.45, 0.72) (*11*). For round 2,the halving time was 8.7 (5.3, 22) days and R was 0.58 (0.37, 0.83). The trends in prevalence that best fit rounds 1 and 2 individually suggest a small increase in prevalence during the unobserved period between rounds (Figure S2).

We compared our estimated trends for national prevalence with publicly available case data from PHE (Figure S3, Table S4). PHE case data are split into health care associated tests (Pillar 1) and tests requested by people with symptoms in the community (Pillar 2). The rate of decline in PHE Pillar 2 case data was similar to that observed when the data from rounds 1 and 2 of REACT were analysed jointly.

Of the 111 participants who tested positive with known symptom status in the week prior to their swab, 21 in our community sample reported symptoms while 90 did not, giving an estimate that 81% (73%, 87%) of people were symptom-free for the 7 days prior to testing positive. This is an increase (p=0.04) from 69% (61%, 76%) in round 1 which may suggest that some symptomatic individuals took advantage of the additional availability of testing from PHE and therefore did not take part in our study. Nonetheless, the high proportion of swab-positive individuals not reporting recent symptoms in both rounds 1 and 2 would be missed by symptom-based surveillance and therefore are a potential source of ongoing transmission that would escape case-based interventions.

We conducted sensitivity analyses to mitigate the risk that the trends we observed from one round to another were biased by systematic differences in the participation of symptomatic individuals or changes in assay sensitivity. We estimated similar growth rates when we restricted our analysis only to asymptomatic individuals testing positive and positive samples for which both E and N genes were detected (Figure S4, Table S4).

Individual-level data allowed us to compare the characteristics of those infected in round 2 versus round 1. Although health care and care home workers were infected far more frequently than other workers in round 1, the odds were markedly reduced in round 2 (Figure 2, Table S3). There are a number of possible explanations for this finding. Testing of health care workers was increasing during this period (Pillar 1) and so individuals may have opted to be tested elsewhere. However, the proportion of our sample that were health care or care home workers increased from round 1 to round 2 (Table 1). Also, trends were similar among asymptomatic individuals (Figure S4). Therefore, we suggest that this result most likely reflects improvements in infection control in health care (*13*) and care home settings.

**Figure 2.**
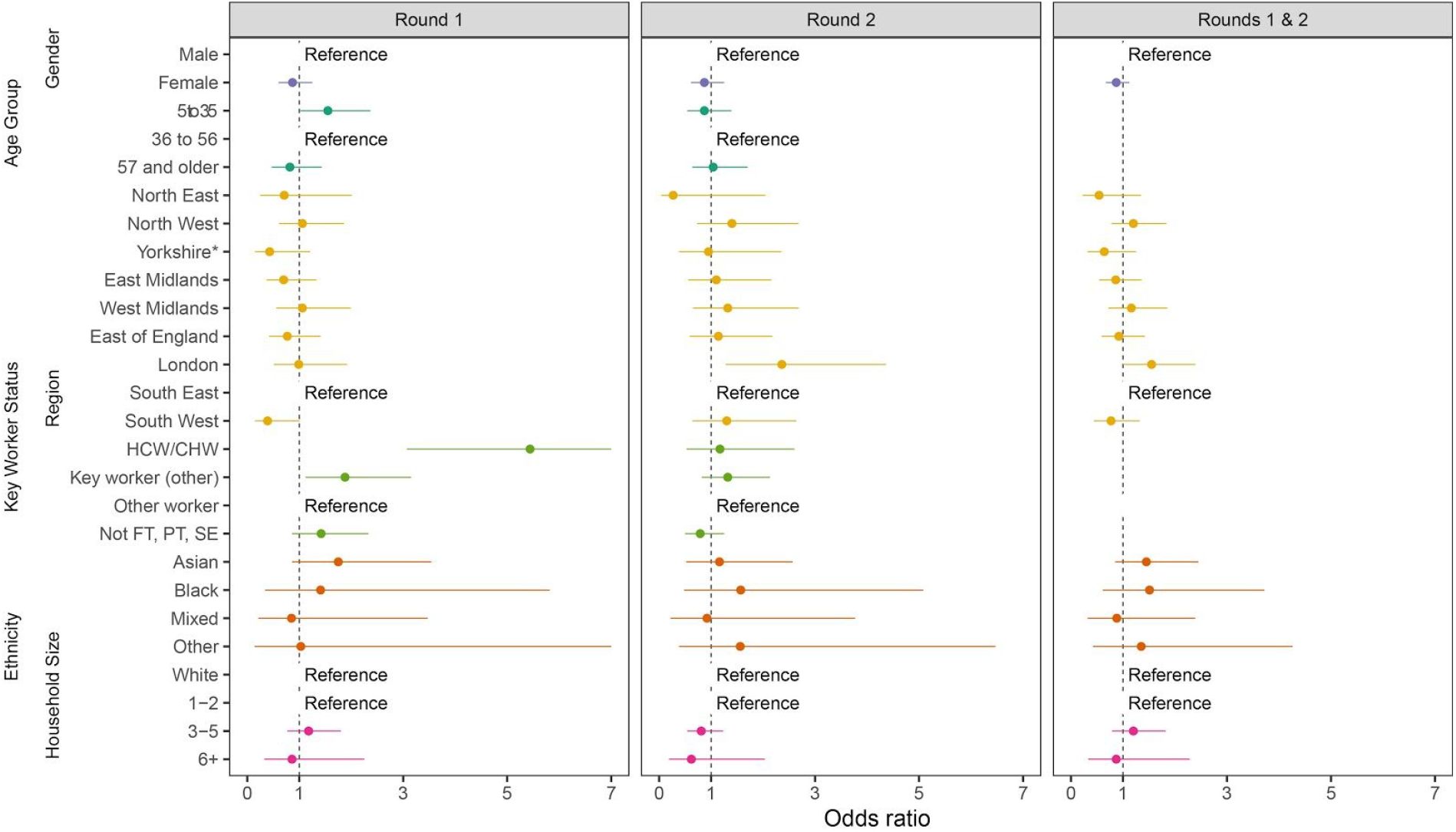
Swab-positivity odds ratios for key epidemiological characteristics. Odds ratios and 95% confidence intervals for epidemiological characteristics (gender, age group, region, key worker status, ethnicity, and household size). Odds ratios were obtained by performing multivariable logistic regression REACT 1 data from round 1, round 2, and rounds 1 and 2 together. The model for rounds 1 and 2 together was jointly adjusted for round, gender, age, region, key worker status, ethnicity, and household size with interaction terms for age by round and key worker status by round. The leftmost plot (Rounds 1 & 2) only shows odds ratios for variables that were fitted without an interaction term. Within the plot, “Reference” indicates the category that was used as the reference group. * Yorkshire and The Humber. HCW / CHW = health care worker / care home worker, Not PT, FT, SE = Not part-time, full-time, or self-employed.

**Table 1.**
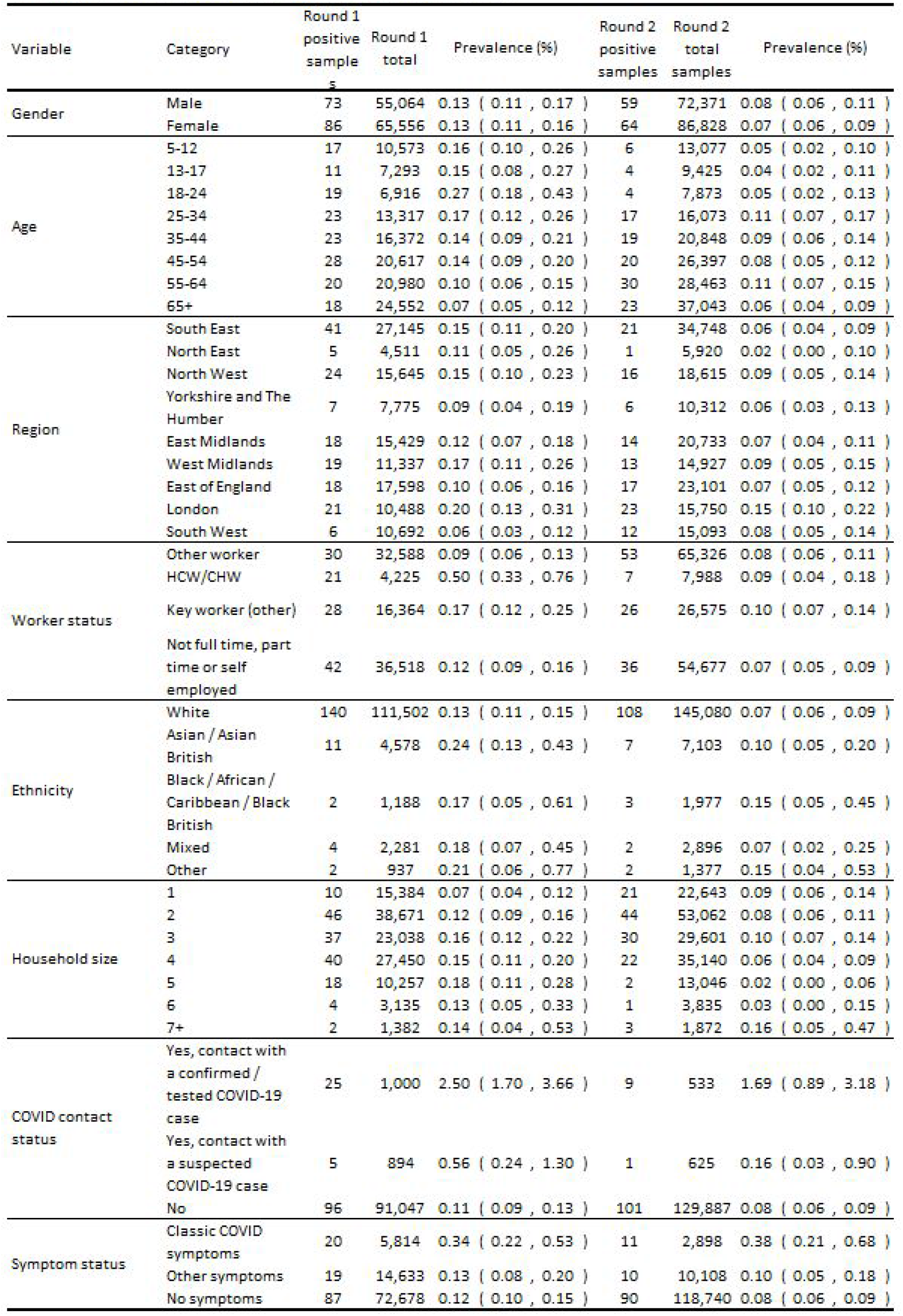
Prevalence cf infection by key characteristic for rounds 1 and 2.

Patterns in unadjusted prevalence suggest differences in social mixing by age between rounds 1 and 2. In round 1 the highest prevalence of 0.27% (0.18%, 0.43%) was found in 18-24 year olds, while children and older adults had similar but lower rates (Table 1). However, in round 2, there was a marked fall in prevalence in 18-24 year olds. Their univariable odds ratio fell from 1.96 (1.1,3.6) in round 1 to 0.56 (0.19, 1.64) in round 2 compared to the 35 to 44 years age group. Although there were also falls in prevalence between rounds in children and adults under the age of 55, for adults aged 55 and over there was little change. These broad patterns by age are maintained in the mutually adjusted regression model (Figure 2, Table S1) indicating that they are not readily explained by measured confounders.

Also, these viral prevalence data were suggestive of increased risk of infection in Black and Asian ethnicities at the end of the lockdown period (Table 1, Figure 2). These observations are consistent with studies of hospitalizations (*14*) and deaths (*15*) showing that these groups are at increased risk of adverse outcomes. Further studies are required to determine the extent to which this increased risk of poor health outcomes can be explained by differential rates of infection.

Of the nine English regions, risk of infection was higher in London than in other areas (Figure 2, Table 1, Table S3). Although the epidemic was geographically diffuse at the time of our study, SARS-CoV-2 transmission first took off in London (*10*). Therefore higher prevalence in London at the end of lockdown suggests that regional rates of decline were similar. This is supported by regionally stratified analysis of our data which did not indicate differences in rates of decline between regions (Figure S5).

We also tested for the presence of clustering at arbitrary spatial scale, independent of geographical units. We compared the distance between all positive pairs and randomly chosen sets of the same number of negative pairs and detected an excess of positive pairs separated by less than 30 km (Figure 3A). We located individual participants who more frequently formed those pairs than would be expected by chance (Figure 3B). Other than one individual in the north west, all participants who were over-represented in nearby pairs lived in or near London (Figure 3C). Conversely, simulated cases from negative pairs were dispersed widely across England (Figure 3D). These analyses suggest that an area of increased infection intensity extended beyond the border of the London geographical region.

**Figure 3.**
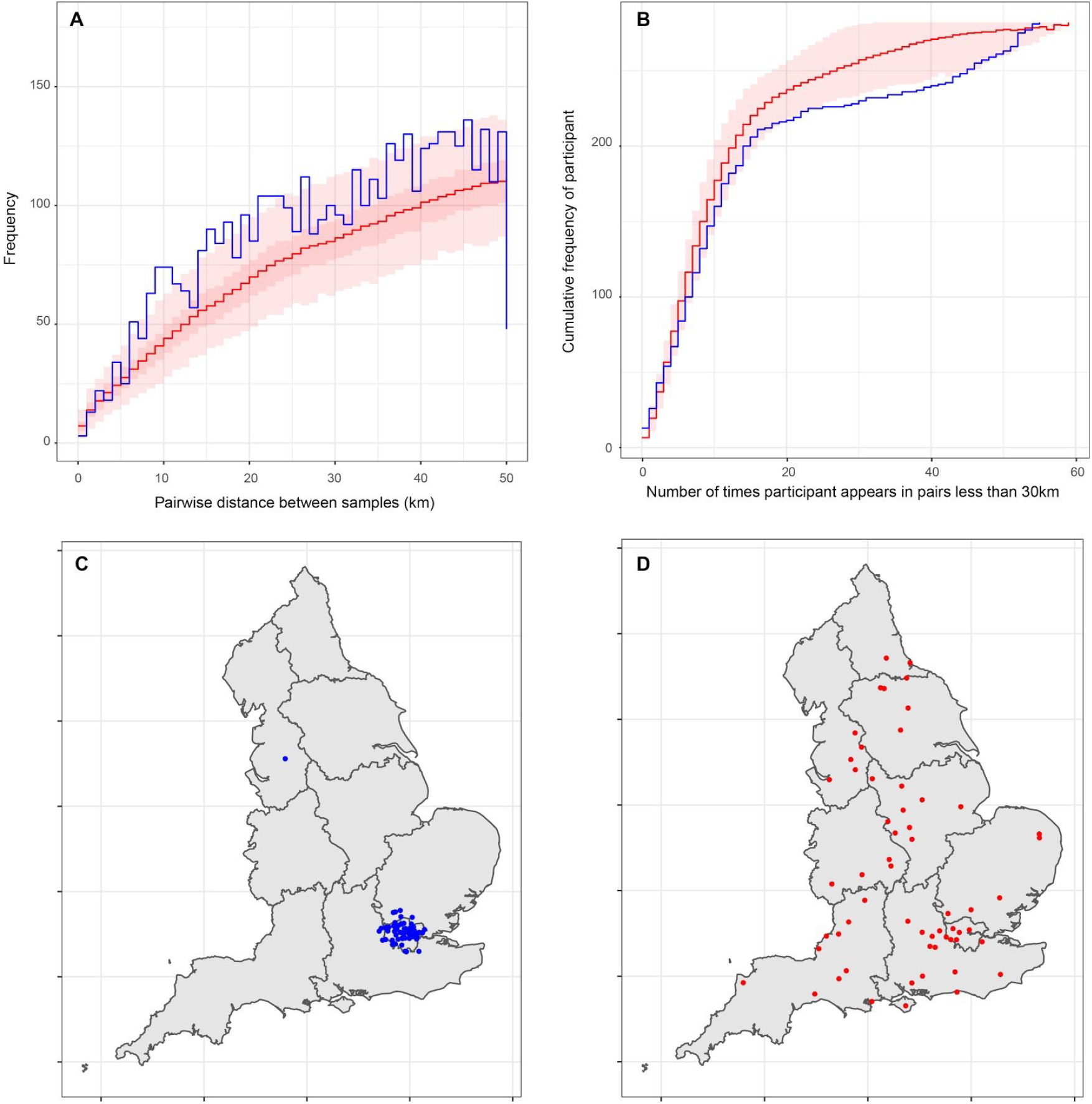
Detection of spatial clustering in and near London for rounds 1 and 2. **A** comparison of the pairwise distance up to 50 km between the home addresses of participants with positive samples for rounds 1 and 2 (n=282, blue) and 1,000 random samples of the same number of home addresses of participants with negative samples (red, mean solid line, central 50% range dark red, full range light red). **B** Cumulative distribution of the number of times participants with positive (blue) or negative (red) tests are in pairs of distance 30 km apart or less. Red line averaged over 1000 random draws of 282 negative participants. Red area is the central 95% range. **C** map of (jittered) location of positive participants who were in 26 or more positive pairs less than 30 km (n=55) **D** map of (jittered) location of participants with negative tests who were in 26 or more negative pairs less than 30 km. Multiple draws of sets of 282 negative participants were needed to generate 55 control locations.

## Conclusion

As the SARS-CoV-2 epidemic in England transitioned out of its initial lockdown phase, prevalence of swab-positivity continued to decrease. The current primary control strategy in the UK is to test, trace and isolate cases and to monitor local case incidence closely so that any additional restrictions on social interactions are conducted at a local level (*16*). Community-based as well as symptomatic testing is essential to inform these strategies.

## Data Availability

The original datasets generated or analysed, or both, during this study are not publicly
available because of governance restrictions and the identifiable nature of the data.

## Contributors

SR and PE conceptualised and designed the study and drafted the manuscript. SR, KECA, OE, CEW and HW undertook the data analysis. PJD, DA and CAD provided statistical advice. GC, WB, HW, CA and AD provided study oversight. AD and PE obtained funding. SR, KECA, OE, CEW, HW, CA, PJD, DA, CAD, GC, WB, HW, GT, AD and PE critically reviewed the manuscript. All authors read and approved the final version of the manuscript. PE is the guarantor for this paper.

## Declaration of interests

We declare no competing interests.

## Funding

The study was funded by the Department of Health and Social Care in England.

## Acknowledgements

SR acknowledges support: MRC Centre for Global Infectious Disease Analysis, National Institute for Health Research (NIHR) Health Protection Research Unit (HPRU), Wellcome Trust (200861/Z/16/Z, 200187/Z/15/Z), and Centres for Disease Control and Prevention (US, U01CK0005-01-02). GC is supported by an NIHR Professorship. PE is Director of the MRC Centre for Environment and Health (MR/L01341X/1, MR/S019669/1). PE acknowledges support from the NIHR Imperial Biomedical Research Centre and the NIHR HPRUs in Environmental Exposures and Health and Chemical and Radiation Threats and Hazards, the British Heart Foundation Centre for Research Excellence at Imperial College London (RE/18/4/34215) and the UK Dementia Research Institute at Imperial (MC_PC_17114).

We thank key collaborators on this work -- Ipsos MORI: Kelly Beaver, Sam Clemens, Gary Welch, Andrew Cleary and Kelly Ward; Institute of Global Health Innovation at Imperial College: Gianluca Fontana, Dr Hutan Ashrafian, Sutha Satkunarajah and Lenny Naar; MRC Centre for Environment and Health, Imperial College London: Daniela Fecht; North West London Pathology and Public Health England for help in calibration of the laboratory analyses; NHS Digital for access to the NHS register; and the Department of Health and Social Care for logistic support. SR acknowledges helpful discussion with members of the UK Government Office for Science (GO-Science) Scientific Pandemic Influenza - Modelling (SPI-M) committee.

## Supporting material for: Transient dynamics of SARS-CoV-2 as England exited national lockdown

**Table S1.** Positive and negative outcomes for rounds 1 and 2 by laboratory and gene detection status.

| Description | Outcome | Round 1 | Round 2 |
| --- | --- | --- | --- |
| PHE, negative result | Negative | 8,579 | 0 |
| PHE, positive result | Positive | 16 | 0 |
| Commerical, no signal for either gene | Negative | 111,437 | 158,447 |
| Commerical, signal for both genes | Positive | 69 | 65 |
| Commercial, signal E gene, no signal N gene | NA | 0 | 0 |
| Commerical, no signal E gene, signal for N gene, CT < 37 | Positive | 74 | 58 |
| Commercial, no signal E gene, signal for N gene, CT ≥ 37 | Negative | 445 | 629 |
| Total number of swabs for which a result is available | - | 120,620 | 159,199 |

**Table S2.** Comparison of parsimony scores for temporal regression models fit to national and regional data (Models fit to both data sets are shown in Figure S5).

| Rounds from<br>which data are<br>used | National / Region | AIC |  |  |
| --- | --- | --- | --- | --- |
|  |  | Smooth term | Linear | Difference |
| Both | National | 4272 | 4292 | -20.7 |
|  | North East | 101 | 101 | 0.0 |
|  | North West | 586 | 588 | -2.0 |
|  | Yorkshire and The Humber | 217 | 217 | 0.0 |
|  | East Midlands | 509 | 510 | -1.3 |
|  | West Midlands | 463 | 463 | 0.0 |
|  | East of England | 549 | 549 | -0.3 |
|  | London | 621 | 623 | -2.2 |
|  | South East | 950 | 950 | 0.0 |
|  | South West | 293 | 300 | -7.3 |
| Round1 | National | 2274 | 2277 | -3.0 |
|  | North East | 81 | 81 | 0.0 |
|  | North West | 328 | 328 | 0.0 |
|  | Yorkshire and The Humber | 115 | 115 | 0.0 |
|  | East Midlands | 274 | 274 | 0.0 |
|  | West Midlands | 253 | 254 | -1.5 |
|  | East of England | 267 | 267 | 0.0 |
|  | London | 271 | 271 | 0.0 |
|  | South East | 597 | 597 | -0.5 |
|  | South West | 103 | 104 | -0.8 |
| Round2 | National | 1994 | 1999 | -5.5 |
|  | North East | 23 | 23 | 0.0 |
|  | North West | 257 | 257 | 0.0 |
|  | Yorkshire and The Humber | 105 | 105 | -0.1 |
|  | East Midlands | 235 | 235 | 0.0 |
|  | West Midlands | 211 | 211 | 0.0 |
|  | East of England | 283 | 283 | 0.0 |
|  | London | 349 | 349 | 0.0 |
|  | South East | 354 | 356 | -1.2 |
|  | South West | 189 | 189 | 0.0 |

**Table S3.** Logistic regression models of swab-positivity as a function of gender, age, region, worker status, ethnicity and household size.

| Characteristic | Category | Round 1 |  |  | Round 2 |  |  | Rounds 1 and 2<br>Fully adjusted, with interactions |  |  |  |
| --- | --- | --- | --- | --- | --- | --- | --- | --- | --- | --- | --- |
|  |  | Unadjusted | Adjusted for age and sex | Fully adjusted | Unadjusted | Adjusted for age and sex | Fully adjusted | Unadjusted | Adjusted for age and sex | Round 1 | Round 2 |
| Gender | Male | Ref | Ref | Ref | Ref | Ref | Ref | Ref | Ref | Ref | Ref |
|  | Female | 0.99 [0.72,1.35] | 0.98 [0.71,1.33] | 0.87 [0.60,1.25] | 0.90 [0.63,1.29] | 0.90 [0.63,1.29] | 0.87 [0.61,1.25] | 0.95 [0.75,1.20] | 0.94 [0.75,1.19] | 0.87 [0.67,1.12] |  |
| Age | 5 to 35 | 1.66 [1.15,2.38] | 1.66 [1.15,2.38] | 1.55 [1.02,2.37] | 0.79 [0.50,1.25] | 0.79 [0.50,1.24] | 0.87 [0.54,1.39] | 1.26 [0.95,1.66] | 1.26 [0.95,1.66] | 1.54 [1.01,2.35] | 0.88 [0.55,1.40] |
|  | 36 to 56 | Ref | Ref | Ref | Ref | Ref | Ref | Ref | Ref | Ref | Ref |
|  | 57 or older | 0.73 [0.47,1.13] | 0.73 [0.47,1.13] | 0.82 [0.47,1.43] | 0.96 [0.64,1.45] | 0.96 [0.64,1.45] | 1.04 [0.64,1.70] | 0.84 [0.62,1.13] | 0.84 [0.62,1.13] | 0.83 [0.48,1.44] | 1.03 [0.63,1.68] |
| Region | North East | 0.73 [0.29,1.86] | 0.74 [0.29,1.87] | 0.71 [0.25,2.01] | 0.28 [0.04,2.08] | 0.28 [0.04,2.07] | 0.27 [0.04,2.04] | 0.57 [0.25,1.33] | 0.58 [0.25,1.33] | 0.54 [0.22,1.35] |  |
|  | North West | 1.02 [0.61,1.68] | 1.01 [0.61,1.68] | 1.06 [0.61,1.86] | 1.42 [0.74,2.73] | 1.42 [0.74,2.73] | 1.40 [0.73,2.68] | 1.17 [0.78,1.74] | 1.17 [0.78,1.73] | 1.20 [0.78,1.83] |  |
|  | Yorkshire and the Humber | 0.60 [0.27,1.33] | 0.60 [0.27,1.34] | 0.43 [0.15,1.21] | 0.96 [0.39,2.39] | 0.96 [0.39,2.38] | 0.95 [0.38,2.35] | 0.72 [0.39,1.30] | 0.72 [0.40,1.31] | 0.64 [0.32,1.25] |  |
|  | East Midlands | 0.77 [0.44,1.34] | 0.77 [0.44,1.34] | 0.70 [0.37,1.33] | 1.12 [0.57,2.20] | 1.12 [0.57,2.20] | 1.10 [0.56,2.16] | 0.88 [0.58,1.35] | 0.88 [0.58,1.35] | 0.86 [0.54,1.36] |  |
|  | West Midlands | 1.11 [0.64,1.91] | 1.10 [0.64,1.90] | 1.06 [0.56,1.99] | 1.44 [0.72,2.88] | 1.44 [0.72,2.88] | 1.32 [0.65,2.68] | 1.22 [0.79,1.86] | 1.21 [0.79,1.86] | 1.16 [0.72,1.85] |  |
|  | East of England | 0.68 [0.39,1.18] | 0.67 [0.39,1.17] | 0.77 [0.42,1.41] | 1.22 [0.64,2.31] | 1.22 [0.64,2.31] | 1.14 [0.59,2.18] | 0.86 [0.57,1.30] | 0.86 [0.57,1.30] | 0.92 [0.59,1.42] |  |
|  | South East | Ref | Ref | Ref | Ref | Ref | Ref | Ref | Ref | Ref |  |
|  | London | 1.33 [0.78,2.25] | 1.24 [0.73,2.11] | 0.99 [0.51,1.92] | 2.42 [1.34,4.37] | 2.47 [1.37,4.48] | 2.36 [1.28,4.36] | 1.68 [1.14,2.47] | 1.62 [1.10,2.39] | 1.55 [1.00,2.39] |  |
|  | South West | 0.37 [0.16,0.87] | 0.38 [0.16,0.89] | 0.39 [0.15,1.01] | 1.32 [0.65,2.68] | 1.31 [0.64,2.67] | 1.30 [0.64,2.64] | 0.70 [0.41,1.18] | 0.70 [0.42,1.19] | 0.77 [0.44,1.32] |  |
| Worker status | Health care and social care worker | 5.42 [3.10,9.48] | 5.49 [3.11,9.69] | 5.44 [3.07,9.64] | 1.08 [0.49,2.38] | 1.13 [0.51,2.50] | 1.17 [0.53,2.60] | 2.71 [1.76,4.16] | 2.76 [1.79,4.26] | 5.53 [3.14,9.73] | 1.15 [0.52,2.56] |
|  | Other key worker | 1.86 [1.11,3.11] | 1.83 [1.09,3.06] | 1.88 [1.12,3.15] | 1.21 [0.75,1.93] | 1.23 [0.76,1.97] | 1.32 [0.82,2.13] | 1.48 [1.05,2.09] | 1.47 [1.04,2.08] | 1.91 [1.14,3.20] | 1.30 [0.81,2.09] |
|  | Other worker | Ref | Ref | Ref | Ref | Ref | Ref | Ref | Ref | Ref | Ref |
|  | Not full time, part time or self employed | 1.25 [0.78,2.00] | 1.40 [0.86,2.30] | 1.42 [0.86,2.33] | 0.81 [0.53,1.24] | 0.79 [0.50,1.25] | 0.79 [0.50,1.25] | 1.01 [0.74,1.37] | 1.06 [0.76,1.47] | 1.44 [0.88,2.36] | 0.79 [0.50,1.24] |
| Ethnicity | White | Ref | Ref | Ref | Ref | Ref | Ref | Ref | Ref | Ref | Ref |
|  | Asian / Asian British | 1.92 [1.04,3.54] | 1.71 [0.92,3.17] | 1.75 [0.86,3.54] | 1.32 [0.62,2.85] | 1.37 [0.64,2.97] | 1.16 [0.52,2.57] | 1.60 [0.99,2.57] | 1.50 [0.93,2.42] | 1.45 [0.85,2.45] |  |
|  | Black / African / Caribbean / Black British | 1.34 [0.33,5.42] | 1.26 [0.31,5.09] | 1.41 [0.34,5.82] | 2.04 [0.65,6.43] | 2.09 [0.66,6.59] | 1.57 [0.48,5.08] | 1.64 [0.67,3.97] | 1.57 [0.65,3.81] | 1.51 [0.61,3.72] |  |
|  | Mixed | 1.40 [0.52,3.78] | 1.10 [0.40,2.99] | 0.85 [0.21,3.47] | 0.93 [0.23,3.76] | 1.02 [0.25,4.19] | 0.92 [0.22,3.77] | 1.20 [0.53,2.70] | 1.06 [0.47,2.41] | 0.88 [0.32,2.39] |  |
|  | Other | 1.70 [0.42,6.88] | 1.63 [0.40,6.58] | 1.03 [0.14,7.48] | 1.95 [0.48,7.92] | 1.99 [0.49,8.08] | 1.56 [0.38,6.47] | 1.79 [0.67,4.81] | 1.73 [0.64,4.66] | 1.35 [0.42,4.26] |  |
| Household size | 1 or 2 | Ref | Ref | Ref | Ref | Ref | Ref | Ref | Ref | Ref | Ref |
|  | 3 to 5 | 1.51 [1.09,2.10] | 1.14 [0.78,1.66] | 1.18 [0.77,1.80] | 0.81 [0.56,1.16] | 0.80 [0.53,1.22] | 0.81 [0.54,1.23] | 1.15 [0.91,1.47] | 0.97 [0.74,1.29] | 1.20 [0.79,1.82] | 0.80 [0.53,1.21] |
|  | 6 or greater | 1.33 [0.63,2.79] | 0.88 [0.40,1.90] | 0.86 [0.33,2.25] | 0.82 [0.30,2.24] | 0.83 [0.29,2.36] | 0.62 [0.19,2.03] | 1.12 [0.62,2.02] | 0.89 [0.48,1.65] | 0.87 [0.33,2.28] | 0.60 [0.18,1.98] |

**Table S4.**
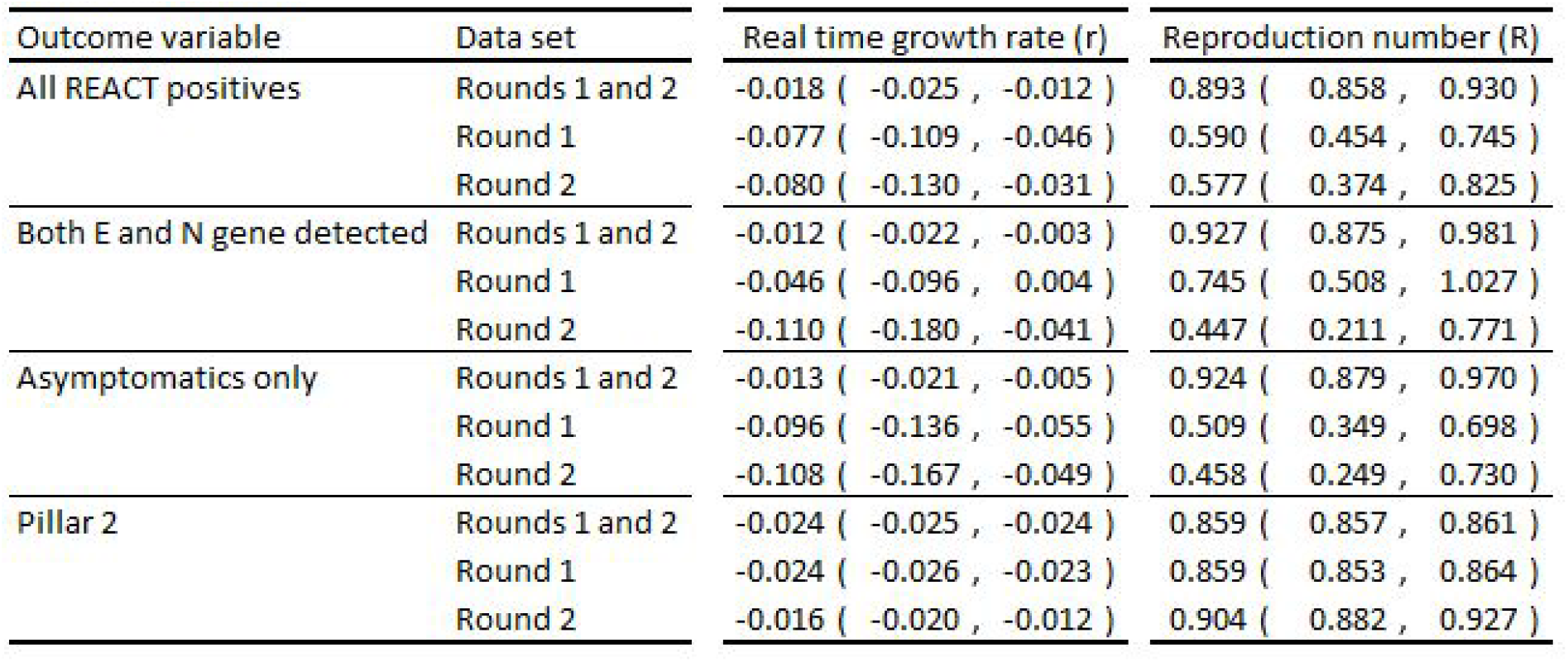
Comparison of estimates of real time growth rate and reproduction number for alternate outcome variables.

**Figure S1.**
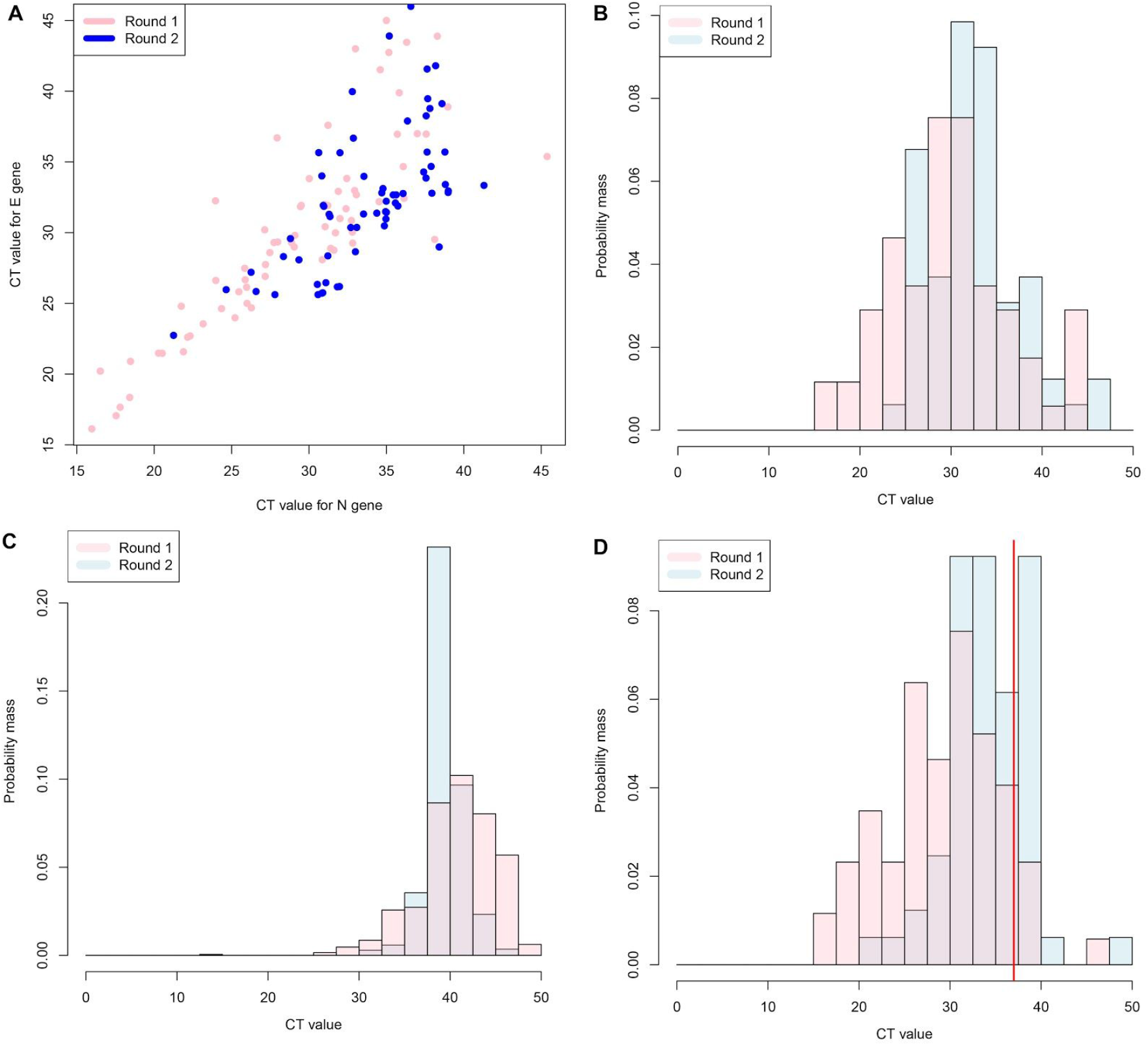
Distributions of cycle threshold values (CT) for rounds 1 and 2. **A** distribution of CT values for samples for which both E and N were detected during round 1 (pink, n=69) and round 2 (blue, n=65). **B** distribution of CT values for E gene for rounds 1 and 2. **C** distribution of CT values for N gene for samples for which both E and N genes were detected. **D** distribution of CT values for N gene for samples for which only the N gene was detected. Vertical red line indicates the positivity threshold of 37.

**Figure S2.**
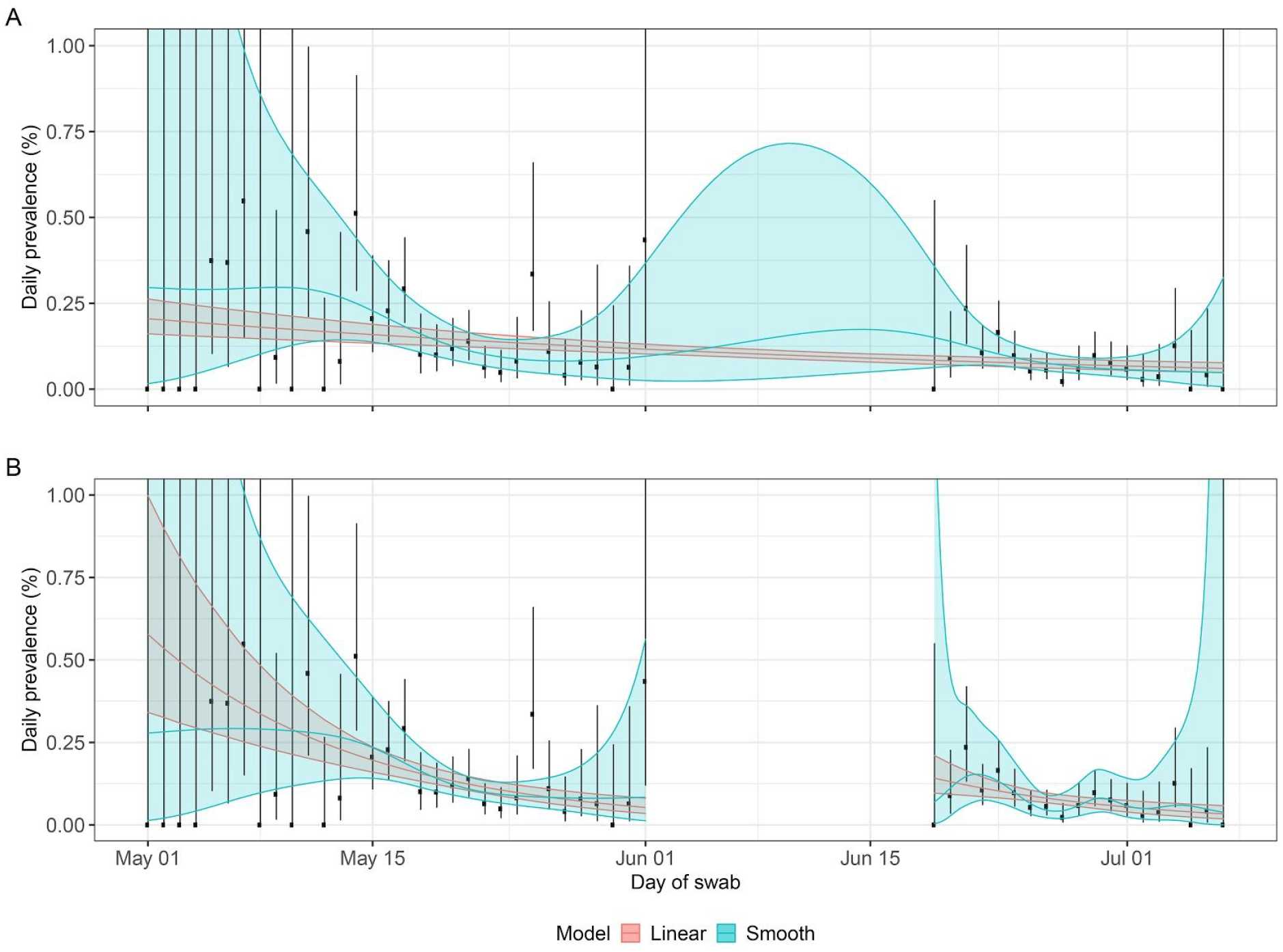
Swab positivity by day with fitted logistic regression models. **A** Observed daily prevalence of swab-positivity for rounds 1 and 2 with regression models fit to both rounds as a single dataset. Red shows best fit for constant decay rate (line) with 95% prediction intervals (shaded area). Blue shows best fit for smooth term decay rate (line) with 95% prediction intervals (shaded area). **B** As A, with models fit separately to rounds 1 and 2.

**Figure S3.**
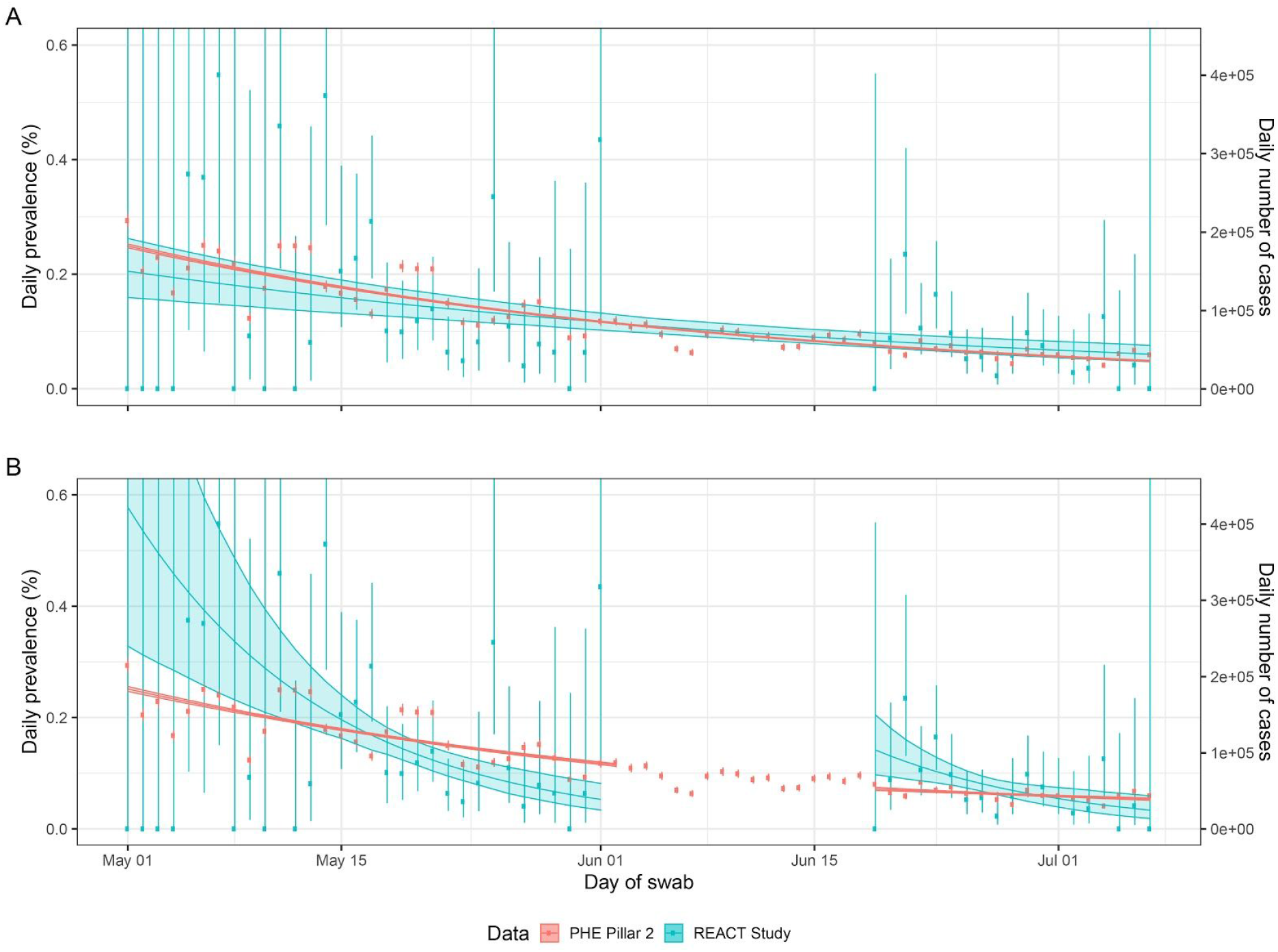
Comparison of fit of exponential model to viral prevalence data from the REACT study rounds 1 and 2 (blue, left y-axis) with Public Health England (PHE, red, right y-axis) non-health care symptomatic testing (Pillar 2) data. **A** single fit (for each data source) to the entire period of the first two rounds of the REACT study. **B** separate fits to the periods of each individual round of the REACT study for both data sources. The left y-axis in A and B is scaled such that PHE cases have the same average vertical location as the REACT study data.

**Figure S4.**
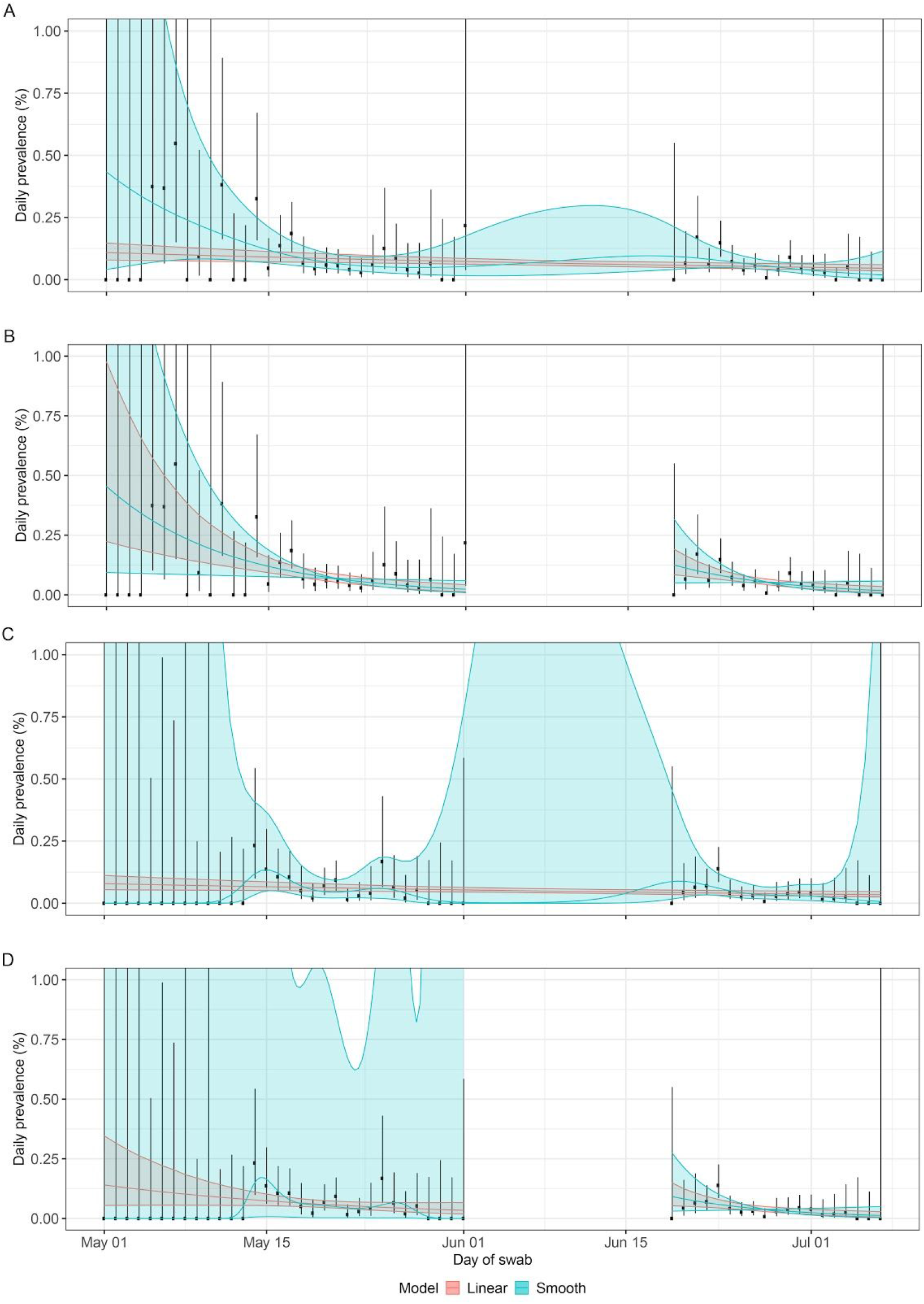
Sensitivity analysis to mitigate risk of false positives. **A** Observed daily prevalence of positive samples from individuals who did not report experiencing symptoms in the week prior to swabbing for rounds 1 and 2 with regression models fit to both rounds as a single dataset. Red shows best fit for constant decay rate (line) with 95% prediction intervals (shaded area). Blue shows best fit for smooth term decay rate (line) with 95% prediction intervals (shaded area). **B** As A, with models fit separately to rounds 1 and 2. **C** Observed daily prevalence of samples for which both E and N genes were detectable for rounds 1 and 2 with regression models fit to both rounds as a single dataset. Red shows best fit for constant decay rate (line) with 95% prediction intervals (shaded area). Blue shows best fit for smooth term decay rate (line) with 95% prediction intervals (shaded area). **D** As C, with models fit separately to rounds 1 and 2. Also see Table S4.

**Figure S5.**
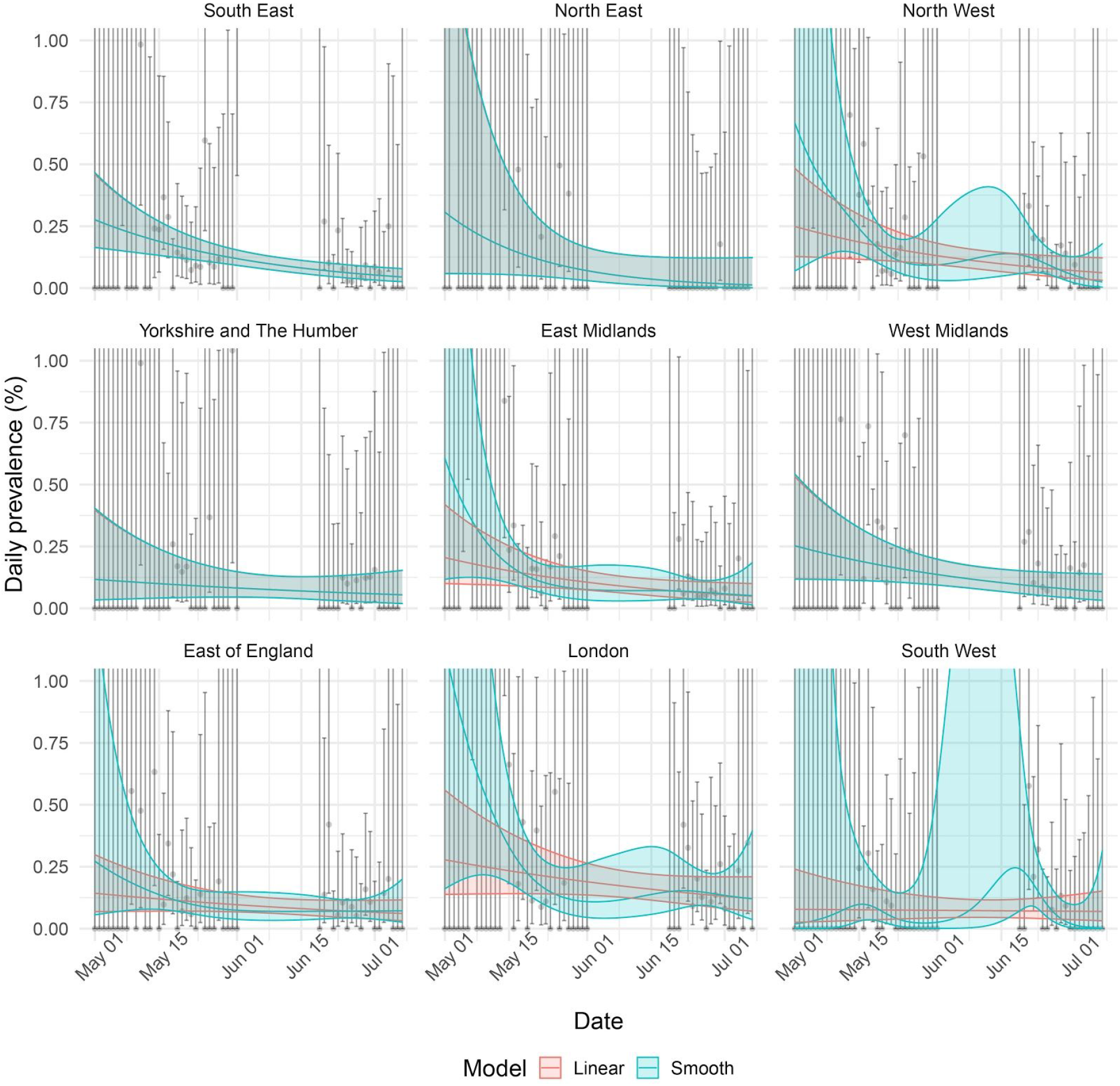
Assessment of temporal trends by region. Observed daily prevalence of swab-positivity for rounds 1 and 2 for each region of England, with regression models fit to both rounds as a single dataset. Red shows best fit for constant decay rate (line) with 95% prediction intervals (shaded area). Blue shows best fit for smooth term decay rate (line) with 95% prediction intervals (shaded area).

## Notes

### Competing Interest Statement

The authors have declared no competing interest.

### Funding Statement

The study was funded by the Department of Health and Social Care in England. SR acknowledges support: MRC Centre for Global Infectious Disease Analysis, National Institute for Health Research (NIHR) Health Protection Research Unit (HPRU), Wellcome Trust (200861/Z/16/Z, 200187/Z/15/Z), and Centres for Disease Control and Prevention (US, U01CK0005-01-02). GC is supported by an NIHR Professorship. PE is Director of the MRC Centre for Environment and Health (MR/L01341X/1, MR/S019669/1). PE acknowledges support from the NIHR Imperial Biomedical Research Centre and the NIHR HPRUs in Environmental Exposures and Health and Chemical and Radiation Threats and Hazards, the British Heart Foundation Centre for Research Excellence at Imperial College London (RE/18/4/34215) and the UK Dementia Research Institute at Imperial (MC_PC_17114).

### Author Declarations

We obtained research ethics approval from the South Central-Berkshire B Research Ethics Committee (IRAS ID: 283787).

## References

1. C. Huang, Y. Wang, X. Li, L. Ren, J. Zhao, Y. Hu, L. Zhang, G. Fan, J. Xu, X. Gu, Z. Cheng, T. Yu, J. Xia, Y. Wei, W. Wu, X. Xie, W. Yin, H. Li, M. Liu, Y. Xiao, H. Gao, L. Guo, J. Xie, G. Wang, R. Jiang, Z. Gao, Q. Jin, J. Wang, B. Cao, Clinical features of patients infected with 2019 novel coronavirus in Wuhan, China. Lancet. 395, 497–506 (2020).

2. J. Oberhammer, Social-distancing effectiveness tracking of the COVID-19 hotspot Stockholm. Epidemiology (2020),, doi:10.1101/2020.06.30.20143487.

3. C. I. Jarvis, K. van Zandvoort, A. Gimma, K. Prem, CMMID COVID-19 working group, P. Klepac, G. James Rubin, W. John Edmunds, Quantifying the impact of physical distance measures on the transmission of COVID-19 in the UK.

4. K. Prem, Y. Liu, T. W. Russell, A. J. Kucharski, R. M. Eggo, N. Davies, Centre for the Mathematical Modelling of Infectious Diseases COVID-19 Working Group, M. Jit, P. Klepac, The effect of control strategies to reduce social mixing on outcomes of the COVID-19 epidemic in Wuhan, China: a modelling study. Lancet Public Health. 5, e261–e270 (2020).

5. E. R. Melnick, J. P. A. Ioannidis, Should governments continue lockdown to slow the spread of covid-19? BMJ. 369, m1924 (2020).

6. K. E. C. Ainslie, C. E. Walters, H. Fu, S. Bhatia, H. Wang, X. Xi, M. Baguelin, S. Bhatt, A. Boonyasiri, O. Boyd, L. Cattarino, C. Ciavarella, Z. Cucunuba, G. Cuomo-Dannenburg, A. Dighe, I. Dorigatti, S. L. van Elsland, R. FitzJohn, K. Gaythorpe, A. C. Ghani, W. Green, A. Hamlet, W. Hinsley, N. Imai, D. Jorgensen, E. Knock, D. Laydon, G. Nedjati-Gilani, L. C. Okell, I. Siveroni, H. A. Thompson, H. J. T. Unwin, R. Verity, M. Vollmer, P. G. T. Walker, Y. Wang, O. J. Watson, C. Whittaker, P. Winskill, C. A. Donnelly, N. M. Ferguson, S. Riley, Evidence of initial success for China exiting COVID-19 social distancing policy after achieving containment. Wellcome Open Res. 5, 81 (2020).

7. B. Jeffrey, C. E. Walters, K. E. C. Ainslie, O. Eales, C. Ciavarella, S. Bhatia, S. Hayes, M. Baguelin, A. Boonyasiri, N. F. Brazeau, G. Cuomo-Dannenburg, R. G. FitzJohn, K. Gaythorpe, W. Green, N. Imai, T. A. Mellan, S. Mishra, P. Nouvellet, H. J. T. Unwin, R. Verity, M. Vollmer, C. Whittaker, N. M. Ferguson, C. A. Donnelly, S. Riley, Anonymised and aggregated crowd level mobility data from mobile phones suggests that initial compliance with COVID-19 social distancing interventions was high and geographically consistent across the UK. Wellcome Open Res. 5, 170 (2020).

8. S. Flaxman, S. Mishra, A. Gandy, H. J. T. Unwin, T. A. Mellan, H. Coupland, C. Whittaker, H. Zhu, T. Berah, J. W. Eaton, M. Monod, Imperial College COVID-19 Response Team, A. C. Ghani, C. A. Donnelly, S. M. Riley, M. A. C. Vollmer, N. M. Ferguson, L. C. Okell, S. Bhatt, Estimating the effects of non-pharmaceutical interventions on COVID-19 in Europe. Nature (2020), doi:10.1038/s41586-020-2405-7.

9. S. Caul, “Deaths registered weekly in England and Wales, provisional: week ending 24 July 2020” (Office of National Statistics, 2020), (available at https://www.ons.gov.uk/peoplepopulationandcommunity/birthsdeathsandmarriages/deaths/bulletins/deathsregisteredweeklyinenglandandwalesprovisional/weekending24july2020).

10. Public Health England, “Weekly Coronavirus Disease 2019 (COVID-19) Surveillance Report. Year 2020. Week 31” (Public Health England, 2020).

11. S. Riley, K. E. C. Ainslie, O. Eales, B. Jeffrey, C. E. Walters, C. J. Atchison, P. J. Diggle, D. Ashby, C. A. Donnelly, G. Cooke, W. Barclay, H. Ward, G. Taylor, A. Darzi, P. Elliott, Community prevalence of SARS-CoV-2 virus in England during May 2020: REACT study. Infectious Diseases (except HIV/AIDS) (2020),, doi:10.1101/2020.07.10.20150524.

12. H. J. Wearing, P. Rohani, M. J. Keeling, Appropriate models for the management of infectious diseases. PLoS Med. 2, e174 (2005).

13. N. K. Jones, L. Rivett, D. Sparkes, S. Forrest, S. Sridhar, J. Young, J. Pereira-Dias, C. Cormie, H. Gill, N. Reynolds, M. Wantoch, M. Routledge, B. Warne, J. Levy, W. D. Córdova Jiménez, F. N. B. Samad, C. McNicholas, M. Ferris, J. Gray, M. Gill, CITIID-NIHR COVID-19 BioResource Collaboration, S. Baker, J. Bradley, G. Dougan, I. Goodfellow, R. Gupta, P. J. Lehner, P. A. Lyons, N. J. Matheson, K. G. Smith, M. E. Torok, M. Toshner, M. D. Curran, S. Fuller, A. Chaudhry, A. Shaw, J. R. Bradley, G. J. Hannon, I. G. Goodfellow, G. Dougan, K. G. Smith, P. J. Lehner, G. Wright, N. J. Matheson, S. Baker, M. P. Weekes, Effective control of SARS-CoV-2 transmission between healthcare workers during a period of diminished community prevalence of COVID-19. Elife. 9 (2020), doi:10.7554/eLife.59391.

14. “Disparities in the risk and outcomes from COVID-19” (Public Health England, 2020), (available at https://assets.publishing.service.gov.uk/government/uploads/system/uploads/attachment_data/file/892085/disparities_review.pdf).

15. R. W. Aldridge, D. Lewer, S. V. Katikireddi, R. Mathur, N. Pathak, R. Burns, E. B. Fragaszy, A. M. Johnson, D. Devakumar, I. Abubakar, A. Hayward, Black, Asian and Minority Ethnic groups in England are at increased risk of death from COVID-19: indirect standardisation of NHS mortality data. Wellcome Open Res. 5, 88 (2020).

16. Department of Health and Social Care, “Breaking chains of COVID-19 transmission to help people return to more normal lives: developing the NHS Test and Trace service” (Department of Health and Social Care, 2020), (available at https://www.gov.uk/government/publications/developing-nhs-test-and-trace-business-plan/breaking-chains-of-covid-19-transmission-to-help-people-return-to-more-normal-lives-developing-the-nhs-test-and-trace-service).

